# The SToP (See, Treat, Prevent) Skin Sores and Scabies Trial - a stepped wedge cluster randomised trial for skin disease control in remote Western Australia: Statistical analysis plan

**DOI:** 10.1101/2023.04.23.23288992

**Authors:** Mark Jones, Hannah Thomas, Thomas Snelling, Asha Bowen, Julie Marsh

**Affiliations:** Faculty of Medicine and Health, University of Sydney, University of Sydney, Sydney, New South Wales, Australia; Wesfarmers Centre for Vaccines and Infectious Diseases, Telethon Kids Institute, University of Western Australia, Perth, Western Australia, Australia

**Keywords:** Skin infections, Australian Aboriginal health, Statistical analysis plan, Stepped wedge cluster design, Bayesian design, Bayesian analysis

## Abstract

The SToP trial is an (open-cohort) stepped-wedge cluster randomised trial (SWCRT) that aims to evaluate a skin health programme comprising three intervention components (1) seeing skin infections; (2) treating skin infections; and (3) preventing skin infections. Four community clusters in the remote Kimberley region of Western Australia will participate in the study. The primary outcome is the diagnosis of impetigo in children (5–9 years) observed during school-based surveillance visits. We provide a detailed, prospective statistical analysis plan (SAP). The plan was written by the trial statistician and details the study design as well as the statistical methods and reporting to be used. The SAP was produced prior to any of the authors viewing any of the trial data. Application of this SAP will minimise bias and supports transparent and reproducible research. SToP is registered under the Australian and New Zealand Clinical Trials Registry, ACTRN12618000520235.

## 1 Background

In remote Australian Aboriginal communities, skin infections (scabies and impetigo) are common. The prevalence for impetigo skin infections has remained largely unchanged over the last decade with approximately 45 % (IQR 34-49%) of children having impetigo at any given time. This translates to almost 16,000 Aboriginal children in remote northern Australia. Despite these concerning statistics, skin infections frequently go untreated due to under-recognition and lack of awareness of their importance. Specifically, skin infections can result in deleterious sequelae such as Streptococcal and Staphylococcal sepsis, bone infections, kidney and rheumatic heart disease.

The SToP trial is an open-cohort stepped wedge cluster randomised trial (SWCRT). This trial has been designed in partnership with Kimberley investigators and will contribute to the ongoing work in the region. The goal is to evaluate an intervention program designed to enhance sustainable skin health practices in remote Aboriginal communities in the Kimberley region of Western Australia. For practical reasons, the intervention components could only be applied at the community level. Under Aboriginal leadership, the study was rolled out in four Kimberley communities, each of which was designated as a cluster. Two of the four community clusters are an amalgamation of communities located in the same vicinity. The intervention components (“SToP activities”) are listed below:

- **Seeing** skin infections through development of training resources/packages within a community dermatology model through school-based surveillance of the primary outcome;
- **Treating** skin infections using the latest evidence implemented using the Structured Administration and Supply Arrangements ‘standing orders’ namely co-trimoxazole for 3 days BD for impetigo, ivermectin on days 0 and 8 for scabies cases and their contacts and holistic care including treatment of those identified with crusted scabies (ivermectin on days 0, 1 and 8 for grade 1 crusted scabies);
- **Preventing** skin infections through embedded, culturally informed and developed health promotion and environmental health activities.

The trial aims to evaluate the impact of the implementation of the intervention components relative to usual care within the community clusters and to estimate the prevalence of impetigo in school aged children between 5 and less than 10 years of age. The expectation is that the intervention will achieve a 50% reduction in the prevalence of impetigo in school aged children between 5 and less than 10 years of age.

## 2 Status

The trial commenced in Apr 2019 and the end date for the study was originally planned to be Nov 2021 with analyses starting in Jan/Feb 2022. However, due to disruptions arising from the COVID 19 pandemic, the trial design was amended and the end date for the study extended to Nov 2022. Follow up is largely complete and at the time of writing, the data is being cleaned by research staff. None of the authors have seen or are privy to any data at the time of writing.

## 3 Study Design

As noted, SToP is an open-label, superiority trial that adopts an open-cohort stepped-wedge cluster randomised trial (SWCRT) design. We introduce some of the characteristics of this type of design in order to orientate the reader.

The SWCRT design is a cluster-based design (randomisation occurs at the cluster-level rather than the individual) with a uni-directional crossover. The intervention is rolled out over time to the different clusters such that a progressively increasing proportion of the study population is exposed. One of the main motivations for creation of the SWCRT was to overcome ethical problems, such as withholding treatments that are already believed to be effective. There are also some statistical benefits, such as the potential for increased power relative to a parallel group cluster design. However, there are a number of challenges associated with the SWCRT.

Figure 1 shows a generic schematic of a hypothetical SWCRT. The schematic illustrates a design with *I* = 8 clusters, *J* = 5 periods and *S* = 4 distinct intervention sequences with two clusters randomised to each sequence. SWCRTs are generally two-armed, i.e. a control vs intervention comparison. Each cluster transitions (steps) from the control state to the intervention state at different timepoints (intervention state denoted by gray shading in figure). All clusters usually start out in the control condition and end in the intervention condition. The discrete periods where observations are made and the interventions switched are usually equally spaced. Study configurations as described are known as ‘complete-designs’ and represent the ideal from a statistical perspective. In contrast, SWCRTs exist where some data points are not collected for a variety of reasons. These are known as ‘incomplete-designs’ and can add further complexity to the modelling/analysis stage.

**Figure 1:**
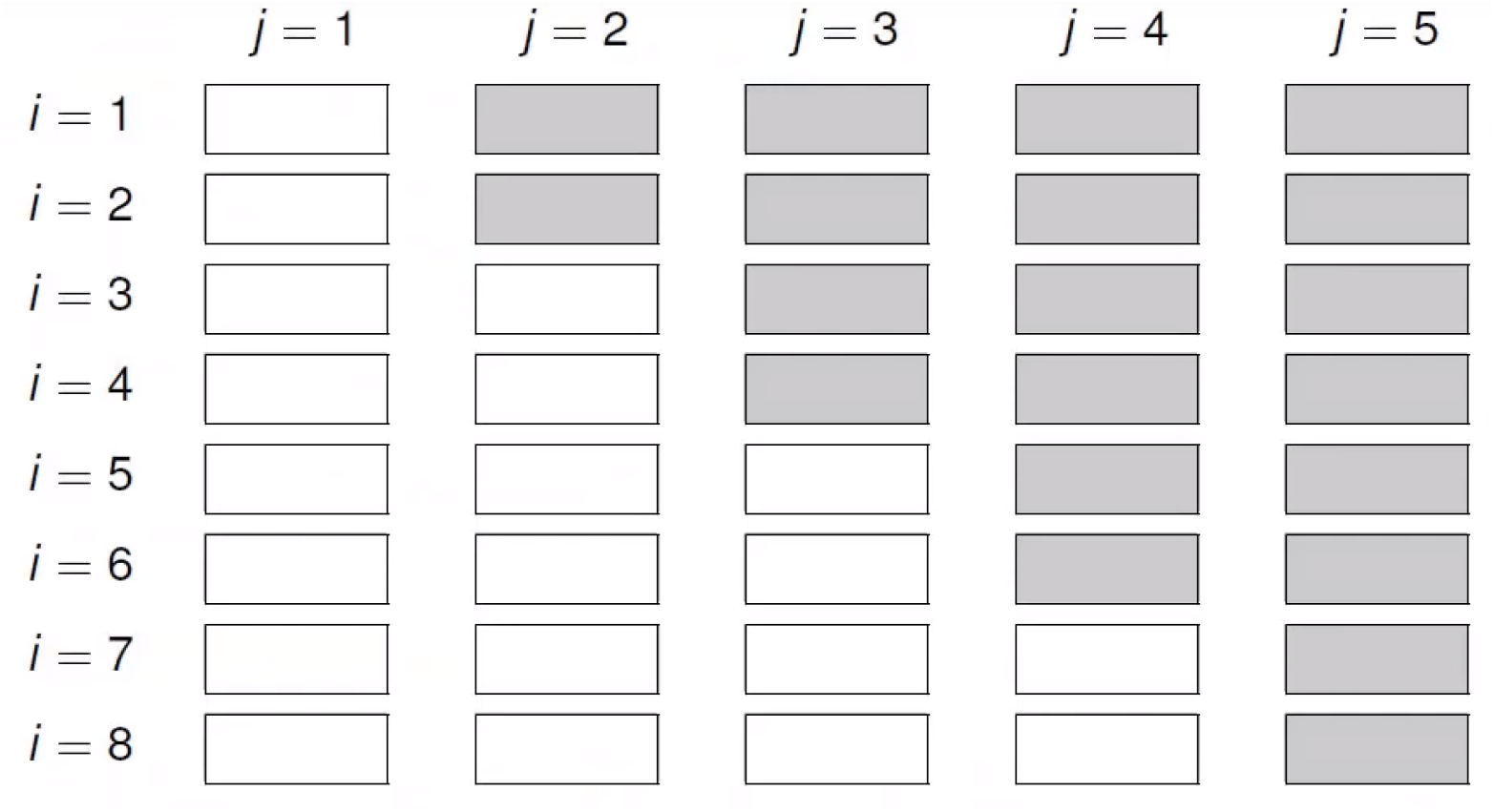
Generic schematic of SWCRT

Despite the novelty and popularity of these designs, SWCRTs are complex and necessitate a range of modelling decisions and assumptions regarding the data generating process (Li et al. 2021). One limitation of the design is that treatment received and time are correlated by design such that the presence of time effects confounds the association between intervention and outcome (temporal-confounding) (Hemming, Taljaard, and Grimshaw 2019). Such a situation can arise in practice when (i) factors external to the study influence the primary outcomes over time, and (ii) the proportion of clusters exposed to the intervention increases with calendar time (always true for the SWCRT design). For example, say that the disease of interest could be monitored via some continuous variable (with a reduction in the metric indicating improvement) derived from the individuals within the cluster and that there was a decline in this metric due to an unrelated public health measure that occurred concurrently with the trial. As time progresses, more observations are made under the treatment condition and this means that, unless time is adequately adjusted for, an effect would be mistakenly attributed to the treatment, even if there were absolutely no treatment effect (Nickless et al. 2018). This can be seen trivially if one imagines the above scenario and consider it in the context of a cross-sectional SWCRT with only a single person observed in each cluster at each time point and then thinking about the average value for the outcome measure by treatment group. Other complexities include changes in correlation structures over time; cluster contamination; time varying treatment effects and design variations (Hemming et al. 2018).

SWCRT designs come in a variety of forms, namely cross-sectional, closed-cohort and open-cohort. In the cross-sectional form, a distinct set of participants are observed at each time period. These are usually the most straight forward of the SWCRTs to analyse. In the closed-cohort form, each cluster starts and ends with the same set of participants that were followed over time and as such the repeat measures on individuals must be accounted for in the analyses. In the open-cohort form, participants enter and leave the clusters, with some being observed contiguously over several periods and others appearing and leaving in a random fashion. Therefore the open-cohort modelling also needs to account for the possibility of repeat measures at the individual level.

SToP is an open-cohort SWCRT and as such we expect to observe some participants multiple times, but other participants will enter and leave the study over its duration. This design was chosen because of the nature of the composite intervention, which is applied at the individual and community-level and the fact that the communities all expressed a wish to receive the intervention. The design for SToP has two distinct sequences, four clusters and was originally planned to have 8 periods. While the SWRCT generally require a greater numbers of clusters, it was not possible to obtain more. The design is incomplete, in that the periods as originally defined were not regularly spaced. This choice of sampling times was adopted to match school term dates and the periods when the remote communities were accessible. However, the irregularity of the sampling points has been further exaggerated due to the impacts of COVID-19. Given the above factors, a cautious approach to the interpretation and generalisation of the results is warranted.

## 4 Intervention

The composite intervention of seeing, treating and preventing will be introduced at the switchover points. Timing, duration and measurement of the intervention is informally described below. Further information on the intervention can be found in the study protocol.

- Timing
  - Groups of individuals are exposed to the intervention.
  - Clinic, school staff and community researchers are exposed to training (see below) at switchover. Refresher training is provided during surveillance activities in subsequent visits.
  - School aged children identified during surveillance as requiring treatment are referred for the treatments defined in the SToP protocol.
  - Community members are exposed to health promotion activities from the consultation phase.

- Duration
  - For each community, training is provided at the start of the intervention switchover for a period of 1 day.
  - The impetigo and scabies treatment regime is in place for the duration of the intervention period for each cluster.
  - Community consultation continues throughout the trial.

- Measurement
  - Repeat measures on school aged children are obtained at the start of surveillance visits.
  - New school aged children may enter at any visit and some children previously observed might not be observed in subsequent periods.

Given the logistical challenges associated with working in remote areas, the intervention components/visits will commence as close as is possible to concurrently.

The control state, which is in place until the switch over points, corresponds to the existing policies and procedures within each community for the diagnosis, treatment and prevention of skin sores and scabies.

## 5 Study population

The trial protocol specifies criteria for both the clusters and participants, specifically: Cluster inclusion criteria:

- Remote Aboriginal community cluster in the Kimberley region of Western Australia
- Community cluster population is around 1,000 people
- Community cluster has access to a clinic staffed full time by nurses
- Community cluster is practically accessible to research staff
- Community cluster indicate interest and consent to participate in trial

Cluster exclusion criteria:

- Community elects not to participate in the trial during consultation phase

All members of consenting communities will be eligible for participation in the SToP trial activities. The inclusion criteria for children participating in ongoing school based surveillance of skin infection rates are:

- Children attending the local community school on the day of surveillance activities
- Informed consent from parent / carer to participate in surveillance activities
- Where feasible, if early childhood education programs exist and are supportive, screening will also occur for the younger age group.

## 6 Sites

The communities themselves were not selected at random but represent a pragmatic sample for which the communities expressed an interest to participate, form distinct geographic clusters and are accessible for trial visits and assessments. Table 1 shows a list of the communities included in the studies, which map to the four clusters.

**Table 1:** Study sites/clusters

| Cluster | Community | School |
| --- | --- | --- |
| 1 | Bidyadanga | La Grange Remote Community School |
| 2 | Djarindjin, Lombadina, Beagle Bay, Ardyaloon | Sacred Heart School, Christ the King Catholic School, One Arm Point remote community school |
| 3 | Balgo, Billiluna, Mulan | Luurnpa Catholic School, Kururrungku Catholic Education Centre, John Pujajangka-Piyirn Catholic School |
| 4 | Warmun | Ngalangangpum school |

Note that while Djarindjin and Lombadina are culturally two separate communities, they are geographically located directly next to each other and share a school and clinic.

## 7 Randomisation

The unit of randomisation in a SWCRT is the cluster. The intervention timing steps were computer randomised to the communities (clusters). The unit of observation are the participant outcomes.

## 8 Outcome Measures

### 8.1 Primary

The primary outcome is diagnosis of impetigo at the school screening sessions (occurring prior to the intervention activities each visit) in children aged 5 and less than 10 years of age as this group have high burden of disease and are readily identifiable. When combined with the number of children screened, this gives the observed prevalence at that point in time. This outcome is likely to reflect improvements in all age classes, but we will be unable to quantify that particular improvement directly.

### 8.2 Secondary

We aim (1) to document the impact of the intervention on other child health indicators, (2) monitor antimicrobial resistance of bacterial skin pathogens with increasing cotrimoxazole use and (3) determine the economic burden of skin infection in school aged children of remote communities and evaluate the cost effectiveness of the trial intervention activities. Only the first two aims are covered in this SAP, and the health economic evaluation and potential changes in the circulating GAS strains will be documented elsewhere.

Inference will focus on:

1. The change in the diagnosis of scabies in school children aged between 5 and less than 10 years.
2. The change in diagnosis of impetigo in children aged 0 - 4 years.
3. The change in diagnosis of scabies in children aged 0 - 4 years.
4. The change in clinic (all-aged) presentations due to skin conditions, including abscess in all age-deciles.
5. The change in all-cause clinic presentations and hospitalisations from the communities, (including those for non-skin e.g. anaemia, skin related e.g. sepsis, and skin causes) in children aged <10 years.
6. Age at first impetigo diagnosis in 12-month cohort after start of intervention (subject to the frequency of impetigo diagnoses)
7. Age at first scabies diagnosis in 12-month cohort after start of intervention (subject to the frequency of scabies diagnoses)
8. Cotrimoxazole resistance in circulating S.aureus and GAS strains (subject to frequencies)
9. Penicillin resistance in circulating S.aureus and GAS strains (subject to frequencies)
10. Methicillin resistance in circulating S.aureus and GAS strains (subject to frequencies)
11. Antimicrobial prescribing for all causes (subject to frequencies)
12. Antimicrobial prescribing for skin infections (subject to frequencies)

### 8.3 Safety and Tolerability

Spontaneous reports of adverse events (unsolicited) will be classified as serious (or not) per protocol definitions.

## 9 Data Sources

Data will be sourced from a custom designed clinical record form (CRF) comprising consent forms, eligibility assessment, visit record, adverse event details and protocol deviations. Data was entered from these sources into a trial database by study personnel (none of the authors were involved in this work).

Community-based diagnoses and prescriptions, i.e. non-school-based, will be obtained from de-identified extracts provided by each clinic using electronic search algorithms based on pre-specified keywords and spelling variations.

All final planned analyses identified in the protocol and SAP will be performed only after the study is completed and the database has been cleaned and locked.

Table 2 provides the scheduled data collection time points. The two right-hand columns give detail on the schedule revisions that arose due to the impacts of COVID-19 and the associated site closures.

**Table 2:** Data collection (site visits)

| Start date of visit | Visit (original) | Original | Visit (updated) | Updated |
| --- | --- | --- | --- | --- |
| 2019-04-01 | 1 | Baseline | 1 | Baseline |
| 2019-07-01 | 2 | Baseline | 2 | Baseline |
| 2019-10-01 | 3 | Treatment starts in clusters 1 and 2 after surveillance and data collection | 3 | Treatment starts in clusters 1 and 2 after surveillance and data collection |
| 2020-04-01 | 4 | Surveillance | - | Site closed, no surveillance |
| 2020-07-01 | 5 | Surveillance | - | Site closed, no surveillance |
| 2020-10-01 | 6 | Treatment starts in cluster 3 and 4 after surveillance and data collection | 4 | Treatment restarts in cluster 1 and 2 after surveillance and data collection |
| 2021-04-01 | 7 | Surveillance | 5 | Surveillance |
| 2021-07-01 | 8 | Surveillance | 6 | Surveillance |
| 2021-10-01 | 9 | Maintenance data collection | 7 | Treatment starts in cluster 3 and 4 after surveillance and data collection |
| 2021-12-01 | - | Trial close | - | - |
| 2022-04-01 | - | - | 8 | Surveillance |
| 2022-07-01 | - | - | 9 | Surveillance |
| 2022-12-01 | - | - | - | Trial close |

Note that site closures were in accordance with WA government policy for remote communities in response to the the emerging COVID-19 health emergency.

## 10 Statistical considerations

The study adopts a Bayesian inference as the mode of analysis. Such an approach is of increasing interest to trialists, but remains under-utilised, partly due to the lack of training and expertise in this area (Zhan et al. 2021; Grantham et al. 2022; Cunanan, Carlin, and Peterson 2016). While Bayesian methods were part of the analysis specification from the outset, they were primarily included as an alternative to deal with potential issues that can arise when using a frequentist approach on such a small number of clusters.

Given the trial disruptions, and after gaining approval from the DSMB, the decision was made to adopt a Bayesian approach throughout. The rationale for this is that we consider the approach to be more appropriate given the context of the trial and the inferential outputs are more accessible than those that would be derived from a frequentist perspective for this trial, which may help with dissemination of the results. Additionally, the repeat-sampling properties of Bayesian approaches can be similar to those of an analogous frequentist approach, as is discussed in a recent simulation study comparing the two (Grantham et al. 2022). As such, the adoption of a Bayesian approach is not considered to have a material impact of the pre-study operating characteristics, which were likely to be very imprecise, again due to the limited number of clusters and the simplifying assumptions that are inherent to the calculations. However, an accompanying supplement provides a small simulation study to evaluate some key operating characteristics.

All analyses described within this SAP will be conducted and/or supervised by trials statisticians (MAJ, JAM) using R, version 4.2.1 or above (R Core Team 2022) and Stan (Stan Development Team 2022). Our reporting aims to comply as closely as is practical to the CONSORT guidelines for stepped wedge trials (Hemming, Taljaard, and Grimshaw 2019). All analyses will be reported in a standalone trial report and subsequently in a main manuscript (and supplementary materials). Any variations from the SAP to be reported in the trial report and publications.

### 10.1 Participant flowchart

Since the previous iteration of the SAP, new CONSORT guidelines have become available (Hemming et al. 2018) and we aim to adhere to these where feasible. Cluster and participant flow diagrams (adapted CONSORT-style) will be used to document the allocations, detailing cluster size, recruitment, loss to follow-up, and missing data (Hemming et al. 2018). Figure 2 provides an indicative representation of the information to be presented in the CONSORT flowchart for STOP, which has 2 sequences and 3 periods (representing baseline, step 1 and step2).

**Figure 2:**
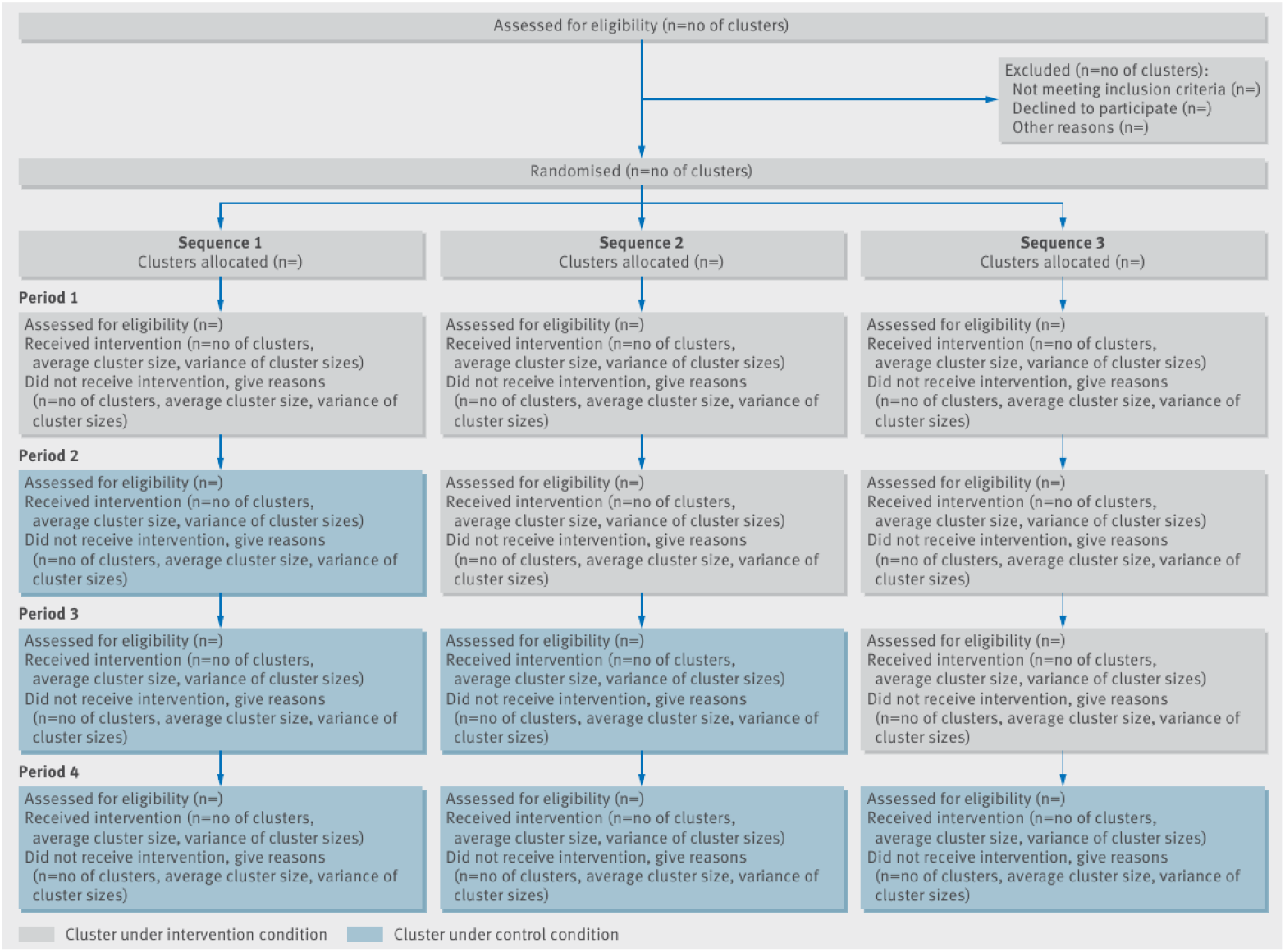
CONSORT - Cluster and participant-level information

### 10.2 Descriptive Statistics - Baseline Characteristics

Community demographics (for the school cohort) and baseline prevalence of impetigo and scabies will be grouped by time of intervention and presented using appropriate summary statistics to assess balance. Figures will be produced to visualise the prevalence of impetigo in each community over time with labelling for the crossover into intervention. We will summarise the following characteristics at baseline data (visit 0 and 1) by cluster and visit:

- number of students surveyed
- weight
- height
- prevalence of impetigo
- prevalence of scabies
- proportion under active treatment for skin infection (past 7 days)
- proportion under any other treatment (past 7 days)
- proportion with visible skin infection
- proportion with skin infection elsewhere on the body (hidden by clothes)
- proportion referred for impetigo and/or scabies/crusted scabies

Categorical variables will be summarised by frequency and percentage. Continuous variables will be summarised by intervention group and overall using median and interquartile range. Missingness will be reported for both categorical and continuous measures.

### 10.3 Descriptive Statistics – Secondary Outcomes

Subject to availability, we will provide descriptive statistics for the secondary outcomes that examine antimicrobial resistance and economic burden. Specifically, we will report:

- Frequency of cotrimoxazole resistance of circulating Staphylococcus aureus and group A strep. (GAS) strains.
- Frequency of antimicrobial prescribing for skin infections and other conditions.

Additionally, subject to availability, we will report:

- Age at first scabies and impetigo diagnosis in the 12-month birth cohort after the SToP activities have been adopted
- Frequency of cotrimoxazole resistance in circulating S. aureus and GAS strains
- Antimicrobial prescribing for skin infections and other conditions

Given the potential impact of the COVID-19 pandemic on the communities, we will provide descriptive statistics for pre-2020 and 2020 for:

- All-cause community clinic presentations
- Diagnosis of skin infections at school screening in children aged 5 and less than 10 years

### 10.4 Analysis of Primary Outcome

The data will be analysed on an intention to treat (ITT) basis with all randomised clusters and observed participants contributing to the primary analysis of the primary endpoint. Contra-indication variations will also be included under ITT.

A generalised linear mixed-effects model (GLMM) framework will be used.

The posterior distribution of each parameter of interest will be reported along with posterior summaries: median, 95% credible interval, and posterior probability of events of interest, e.g. the probability that the treatment effect is less than zero.

### Statistical model notation

Let *i* = 1 … *I* denote the *i*^*th*^ cluster, *j* = 1 … *J* denote the *j*^*th*^ period and *k* = 1 … *K* denote the *kth* participant. Let *yijk* = 1 denote a diagnosis of impetigo for the *kth* participant in the *i*^*th*^ cluster at visit *j* and *yijk* = 0 otherwise. Let *μ* denote the grand mean log-odds of response and let *κ*_*j*_ denote a categorical time effect (secular trend) at each time-step. Let *xij* be an indicator variable for whether cluster *i* is under the intervention state at visit *j* with *δ* representing an average intervention effect, which applies across the whole study period and all clusters. Incorporating within-cluster and within-subject variation (*ν*_*i*_ and *γ*_*ik*_ respectively), we have:

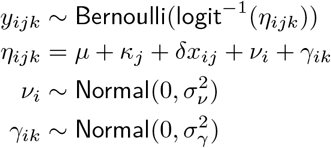

Where the variance terms aim to account for cluster level departure from the overall mean and repeat measures on participants. The above model represents the simplest approach feasible for these data. We note that in its present form, the model does not attempt to characterise treatment effect heterogeneity, nor cluster by time heterogeneity (cluster-specific variation in the secular trend) as these are likely to be weakly informed by the data.

Under a Bayesian analysis, priors must also be specified on all parameters. The priors adopted in the linear predictor have been calibrated to regularise the parameters within plausible ranges. Readers are referred to Gelman, Simpson, and Betancourt (2017), McElreath (2020) and Spiegelhalter, Abrams, and Myles (2004) for further information. We will adopt the following priors noting that the priors are applied on the log-odds scale.

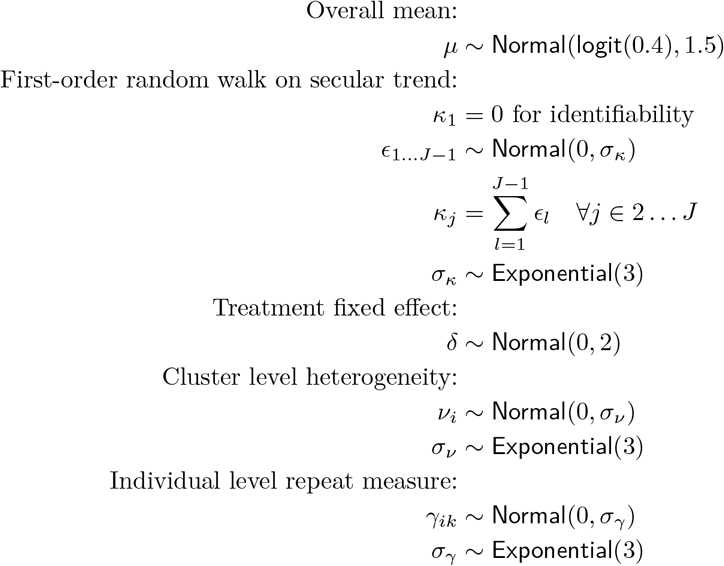

Note - *μ* ∼ Normal(logit(0.4), 1.5) was based on a historical estimate for impetigo prevalence.

Sensitivity analyses will be undertaken via adopting more routinely used priors, e.g. zero centered with large variances in order to examine the sensitivity to the default priors. Further priors may also be used, for example, to investigate the perspective of skeptical a-priori belief in treatment benefit.

Analogous, but not entirely equivalent frequentist models may be run as another mode of model verification.

### 10.5 Analyses of Secondary Outcomes

Table 3 provides a brief summary of the approach to secondary outcome analyses. Secondary analyses will be handled in an analogous manner to the primary analysis with equivalent linear predictors, where applicable. The analyses of health economic data, qualitative surveys, quality of life surveys and changes in circulating GAS strains will be documented separately by appropriate specialists.

**Table 3:** Summary of secondary analyses

| ID | Outcome description | Note | Methods |
| --- | --- | --- | --- |
| 1-CHI | Diagnosis of scabies in children aged between 5 and less than 10 years. | Dichotomous dependent variable with subject level (repeat measure) and community level clustering. Data obtained from school surveillance at each visit. Measure of interest is the treatment effect. | Methods analogous to those used in primary analysis. GLMM (binomial, logit). Random intercept for subject. |
| 2-CHI | Diagnosis of impetigo in children aged between 0 to 4 years. | As above. | As above. Separate model. |
| 3-CHI | Diagnosis of scabies in children aged between 0 to 4 years. | As above. | As above. Separate model. |
| 4-CHI | Overall clinic presentations due to skin conditions including abscesses in all age-deciles. | Analysis pertains to the study period, not beyond. All age classes included. If we have denominators (presumably size of community at each timepoint), we can look at proportion or rate, i.e. incorporate an offset into the model. If no denominators, can look at frequency of visits. Subject and community clustering. Measure of interest is treatment effect. | Methods analogous to those used in primary analysis. Specifics TBD. |
| 5-CHI | All-cause clinic presentations and hospitalisations from the communities, (including those for non-skin e.g. anaemia, skin related e.g. sepsis, and skin causes) in children aged <10 years. | As per 1-CHI. Dichotomous dependent variable (presents to hospital/does not present). | As per 1-CHI. |
| 6-CHI | Age at first scabies and impetigo diagnosis in the 12-month birth cohort after the SToP activities have been adopted. | - | Descriptive statistics. Comparison against historical data (where available). |
| 1-AMR | Frequency of cotrimoxazole resistance of circulating *Staphylococcus aureus* and group A strep. (GAS) strains. | - | Descriptive statistics. Comparison against historical data (where available). |

### 10.6 Analyses of Exploratory Outcomes

A time series analysis may be undertaken to explore the impact of the COVID19 pandemic and the associated public health measures on the prevalence of skin conditions in this cohort. This analysis may be conducted independently from the primary analysis, which will be reported on first.

### 10.7 Analyses of Safety Outcomes

No analyses of safety outcomes have been defined.

SAEs and measures of study conduct and implementation by treatment group will be monitored on a regular basis by a Data and Safety Monitoring Board (DSMB). Adverse events will be summarised for the period before intervention and after the start of intervention by sequence.

### 10.8 Missing Values

Reasons for missing data pertaining to the primary endpoint and secondary endpoints (including withdrawal of consent, loss to follow-up, removal from study due to serious side effects, death, or inability to obtain any laboratory results) will be indicated where available. The quantity of this missing data for each cluster, both prior to and during the intervention period, will be compared.

## 11 Quality Control

The SAP and all statistical analyses will be subject to review by a statistician not involved in the statistical analysis, but familiar with the trial.

Study personnel will perform quality control checks on the final tables, listings and figures to ensure accurate reporting in terms of titles, labels and frequencies or totals.

## 12 Configuration and Formatting Guidelines

An indicative set of tables and figures to be reported are described in the following sections.

### 12.1 Tables & Listings

- The summary statistics for clusters will be displayed in randomisation order with the average cluster size and variation displayed in the column headers.
- Summaries for categorical variables will include only levels with observed data. Percentages corresponding to null categories (cells) will be suppressed.
- All summaries for continuous variables will include: N, median and quartiles 25% and 75% (interquartile range). 95% confidence intervals, coefficient of variation (CV) or %CV may be used as appropriate.

### 12.2 Figures

Legends will be included in all figures with more than one level or category for an explanatory variable. No figures will be created with titles to facilitate flexible use of the figures in reports, manuscripts and presentations; figure titles will be added after the file has been inserted into a WORD, PowerPoint or other such document.

### 12.3 Planned Tables

- Table 1. Demographic and baseline characteristics by cluster
- Table 2. Consent, withdrawals, protocol deviations by cluster
- Table 3. Primary outcome by cluster
- Table 4. Secondary outcomes by cluster
- Table 5. SAEs by sequence and pre-/post-intervention

### 12.4 Planned Figures

- Figure 1. Flowchart of patient progression through the study (CONSORT)
- Figure 2. Stepped wedge design and timing of visits
- Figure 3. Prevalence of impetigo at each study visit by cluster
- Figure 4. Prevalence of scabies at each study visit by cluster

## 13 Declarations

### 13.1 Acknowledgements

The author acknowledges Associate Professor Asha Bowen as the Coordinating Principal Investigator who approved the analysis plan as well as Dr. Julie Marsh, Professor Tom Snelling and Dr. Hannah Thomas, all of whom have been involved in discussions regarding the SToP study and have provided comments on the approach documented herein.

### 13.2 Ethics

The SToP trial protocol is approved by the local medical ethical review committees at the University of Western Australia (Reference RA/4/20/4123), the Child and Adolescent Health Service (Approval number RGS0000000584) at Perth Children’s Hospital and by the Western Australian Aboriginal Health Ethics Committee (Reference number: 819). The study has also been reviewed and approved by the Catholic Education Office (Ref. no. RP2017/57) and Western Australian Department of Education (Ref. no. D18/0281633) for school-based surveillance. The study has also been reviewed and supported by the Kimberley Aboriginal Health Planning Forum Research Subcommittee (Ref. no. 2016-14).

### 13.4 Contributors

Methods MJ. Manuscript written by MJ. Study concept and design were conducted by ACB, JAM. Review and comments from all authors.

## Data Availability

All data produced in the present study are available upon reasonable request to the authors

