## Supplemental material for "The SToP (See, Treat, Prevent) Skin Sores and Scabies Trial - a stepped wedge cluster randomised trial for skin disease control in remote Western Australia: Statistical analysis plan"

Simulation-based analysis of operating characteristics

Mark Jones

Document date: 2023-02-21

### 1 Introduction

Given the disruptions to the trial, which led to modifications in the design and also the decision to move to a Bayesian mode of inference, we provide a small, simulation-based evaluation of operating characteristics. Primarily, we focus on the Bayesian repeat-sampling analogues of type-I error and power. We note that the original sample size calculations were based on a simplifying assumption of a cross-sectional design, which would be inherently more informative than SToP's open cohort design. Additionally, the approach assumed evenly spaced followup, which is not well-aligned with the realised design. Here we make the assumption that the design is a closed-cohort rather than open-cohort and retain the evenly spaced followup. Clearly, these are also a simplifying assumptions, but the closed-cohort perspective is a more realistic representation than assuming that the data are cross-sectional with distinct individuals in each cluster and period of observation.

For the simulation parameters/setup, we retained the original perspective that 84 children would be available per cluster, even though this may not be aligned with what is finally observed in the actual trial. We also looked at the impact of dropping the number of children to 30 per cluster, but did not look at the impact of imbalance. We took the initial background prevalence to be 40% and incorporated a linear secular trend, equating to a 5% drop in the background log-odds of response by the end of the followup. For the intra-cluster correlation coefficients (ICCs) (see next section), we assumed values of 0.01 and 0.05 and for $\rho_3$ (cluster ICC) and we assumed values of 0.15 and 0.45 $\rho_{23}$ (repeat measure ICC). We considered $\rho_3 = 0.01$, $\rho_{23} = 0.15$ to be a low case and $\rho_3 = 0.05$, $\rho_{23} = 0.45$ to be a high correlation case. Treatment effects were evaluated in the range starting with the null case (OR = 1) and extending to an OR that would reduce the background prevalence by 50%, i.e. reduce to 20% under the SToP treatment protocol (OR = 0.375). No loss to follow up was assumed. We acknowledge that these scenarios are arbitrary and undoubtedly are a very crude approximation of reality.

For the procedure to compute power/type-I error, for each simulated trial, we compute the posterior probability that the cluster specific log-odds ratio for the treatment effect is less than zero. We considered a probability exceeding 0.95 to indicate treatment benefit (similar in the dichotomising form of a null hypothesis test) and the proportion of such trials out of those simulated for a given scenario, corresponds to the type-I error (in the null scenario of no treatment effect) and power (in those scenarios where treatment benefit was simulated).

Modelling via MCMC is computationally intensive and so we only ran and evaluated each scenario 400 times. We used priors per those specified in the SAP for the primary analysis model.

#### 2 ICC and variance components

We have variance components to address heterogeneity arising from the clusters and repeat measures on individuals within a cluster. If we assume that the outcome arises from a latent variable following a standard logistic distribution, the error variance can be deemed fixed at $\pi^2/3$. We have definitions for the variance and respective ICCs:

$$\begin{aligned}\sigma^2 &= \sigma^2_\nu + \sigma^2_\gamma + \pi^2/3 \\ \rho_3 &= \frac{\sigma^2_\nu}{\sigma^2} \\ \rho_{23} &= \frac{\sigma^2_\nu + \sigma_\gamma}{\sigma^2}\end{aligned}$$

where

- $\sigma^2$ is the total variance
- $\sigma^2_\nu$ is as defined early, namely the cluster level heterogeneity
- $\sigma^2_\gamma$ is the variance component associated with the repeat measures
- $\rho_3$ is the proportion of the total variance which is due to between cluster variance
- $\rho_{23}$ is the correlation between measurements for the same participant within the same cluster

Rearranging the equation for $\rho_3$ we have:

$$\sigma^2_\nu = \sigma^2 \rho_3$$

Substituting for $\sigma^2_\nu$ in the equation for $\rho_{23}$ we have:

$$\begin{aligned}\rho_{23} &= \frac{\sigma^2 \rho_3 + \sigma^2_\gamma}{\sigma^2} \\ \sigma^2_\gamma &= \sigma^2(\rho_{23} - \rho_3)\end{aligned}$$

Substituting both of the above, for the total variance we get:

$$\begin{aligned}\sigma^2 &= \sigma^2 \rho_3 + \sigma^2(\rho_{23} - \rho_3) + \pi^2/3 \\ \pi^2/3 &= \sigma^2 - \sigma^2 \rho_3 - \sigma^2 \rho_{23} + \sigma^2 \rho_3 \\ \pi^2/3 &= \sigma^2(1 - \rho_{23})\end{aligned}$$

So that the variance components can be defined in terms of:

$$\sigma^2 = \frac{\pi^2/3}{1-\rho_{23}}$$
$$\sigma_\nu^2 = \frac{\pi^2/3}{1-\rho_{23}}(\rho_3)$$
$$\sigma_\gamma^2 = \frac{\pi^2/3}{1-\rho_{23}}(\rho_{23}-\rho_3)$$

From which we can induce target correlations by choosing specific variances in the data generation process for the simulation.

### 3 Simulation results

Figure 1 shows power (represented as a proportion rather than percent) curves for selected scenarios based on the design. Hereafter, the scenarios are referred to as:

- high sample size, low correlation (N 84, ICC 0.01, 0.15)
- high sample size, high correlation (N 84, ICC 0.05, 0.45)
- low sample size, low correlation (N 30, ICC 0.01, 0.15)
- low sample size, high correlation (N 30, ICC 0.05, 0.45)

The results suggest that as the number of participants per cluster reduces and the correlations increase, the power reduces. This is as expected.

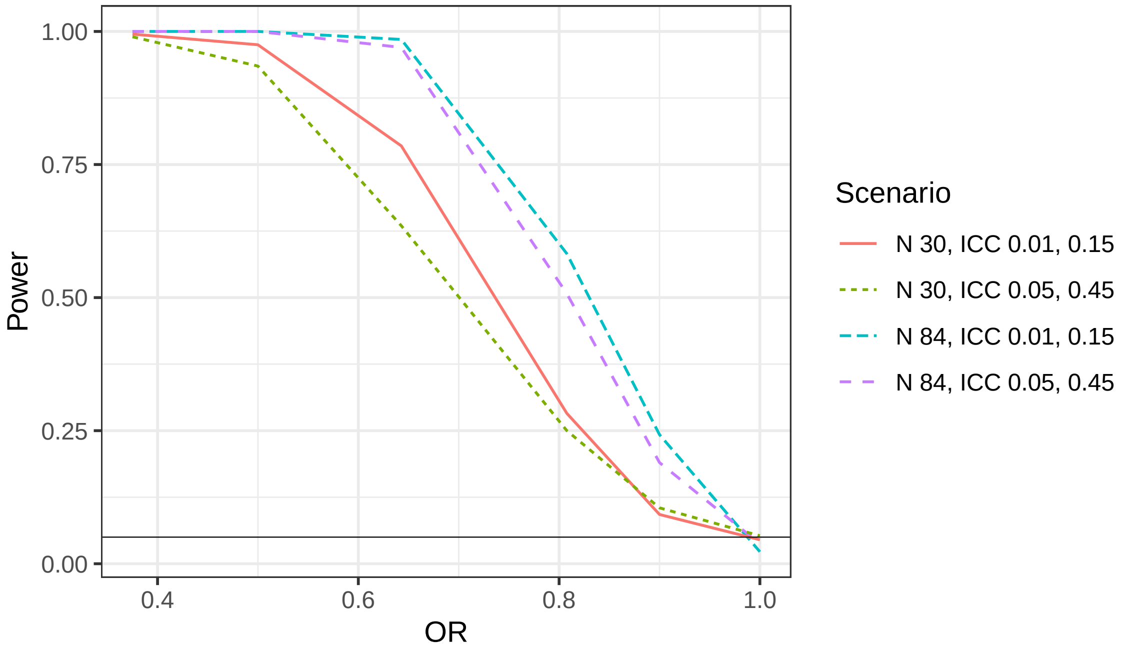


Figure 1: Power curves for selected scenarios

#### 4 Posterior median treatment effect

For each scenario, the true OR and the median of the posterior medians of the OR for the treatment term is shown in figure 2. The vertical lines show the median plus/minus the standard deviation of the posterior median over the simulations. On average, the posterior medians seem reasonable, but there is increasing variance in the point estimates as the number participants decreases and the correlations increase.

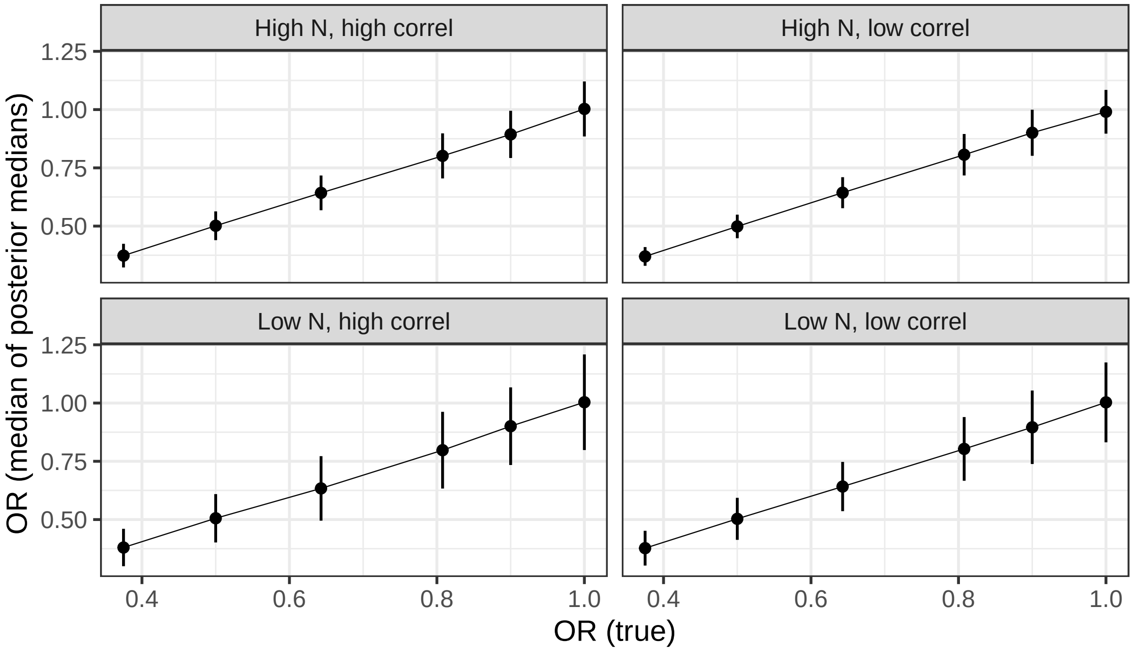


Figure 2: True treatment term OR versus median of posterior medians

### 5 Example results

To give a sense of the assumed prior knowledge, figure 3 shows the prior (dashed) and posterior (solid lines) distribution for the treatment log-OR term obtained from four arbitrary selected datasets generated under the following conditions:

- the null scenario (true OR for treatment effect equal to 1) using high number of participants per cluster
- the null scenario (true OR for treatment effect equal to 1) using low number of participants per cluster
- the positive benefit scenario (true OR 0.64) using high number of participants per cluster
- the positive benefit scenario (true OR 0.64) using low number of participants per cluster

All were based on the low-correlation case (ICC 0.01, 0.15). The vertical lines show the true log-ORs. Some variation from these is to be expected given that the results are based on a single dataset.

Figures 4 and 5 show the analogous output for the variance components (converted into standard deviations). The typical inflation of these parameters can be seen as the posterior is increasingly less informed by the data, i.e. poorer estimation due to less information (specifically less participants).

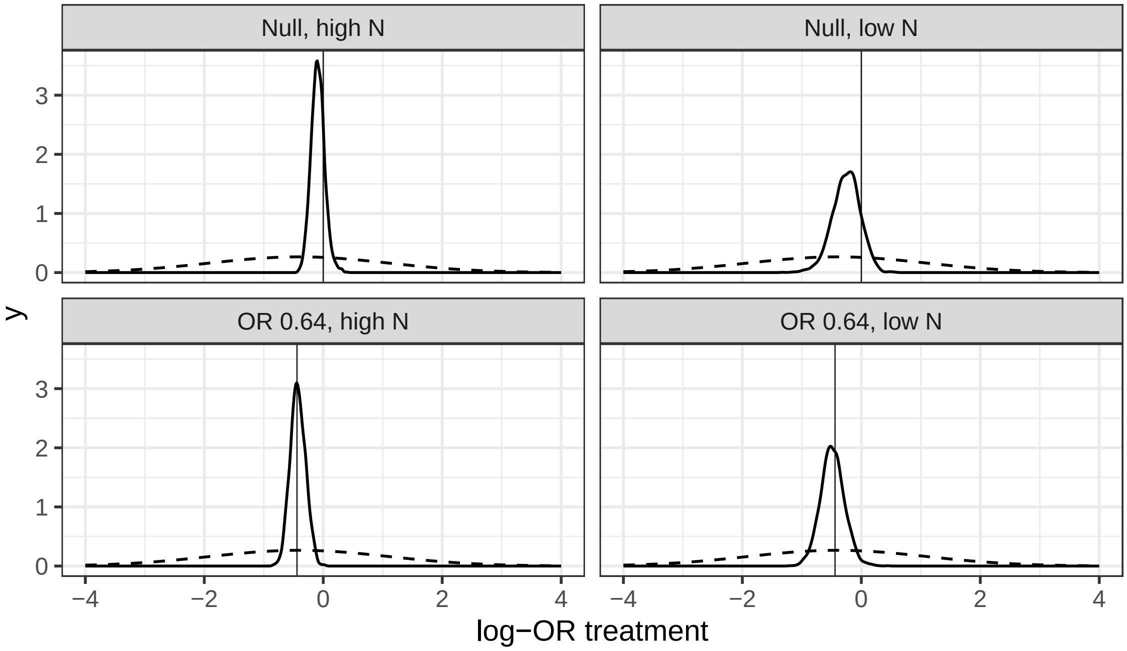


Figure 3: Prior and posterior distributions for treatment log-OR

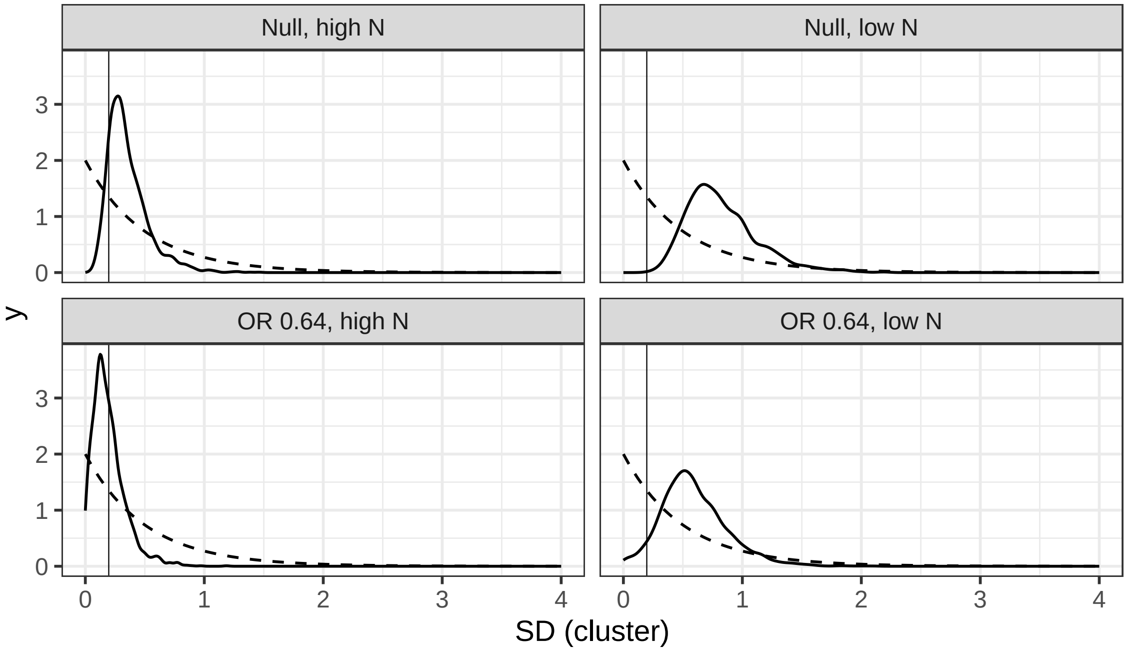


Figure 4: Prior and posterior distributions for variance component on cluster heterogeneity

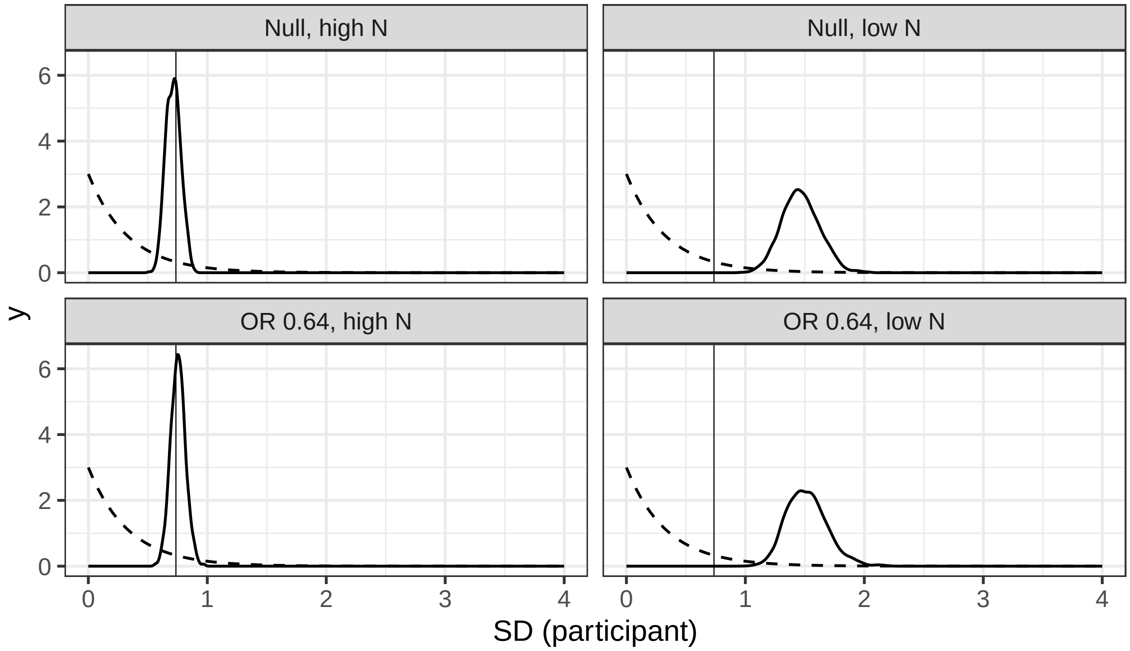


Figure 5: Prior and posterior distributions for variance component on repeat measure
